# Participatory Research: A Priority Setting Partnership for Chronic Musculoskeletal Pain in Denmark

**DOI:** 10.1101/2021.12.17.21267948

**Authors:** KD Lyng, JB Larsen, K Birnie, J Stinson, M Hoegh, AE Olesen, L Arendt-Nielsen, L Ehlers, K Fonager, MB Jensen, H Würtzen, TS Palsson, P Poulin, G Handberg, C Ziegler, LB Møller, J Olsen, L Heise, MS Rathleff

## Abstract

**Background:** Patient and stakeholder engagements in research have increasingly gained attention in healthcare and healthcare-related research. A common and rigorous approach to establish research priorities based on input from people and stakeholders is the James Lind Alliance Priority Setting Partnership (JLA-PSP). The aim of this study was to establish research priorities for chronic musculoskeletal (MSK) pain by engaging with humans living with chronic MSK pain, relatives to humans living with chronic MSK pain, healthcare professionals (HCP), and researchers working with chronic MSK pain.

**Methods:** This JLA-PSP included a nation-wide survey in Denmark, an interim prioritisation, and an online consensus building workshop. The information gained from this was the basis for developing the final list of specific research priorities within chronic MSK pain.

**Results:** In the initial survey, 1010 respondents (91% people living with chronic MSK pain/relatives, 9% HCPs/researchers) submitted 3121 potential questions. These were summarised into 19 main themes and 36 sub-themes. In the interim prioritisation exercise, 51% people living with pain/relatives and 49% HCPs/researchers reduced the list to 33 research questions prior to the final priority setting workshop. 23 participants attended the online workshop (12 people/relatives, 10 HCPs, and 1 researcher) who reached consensus for the most important research priorities after two rounds of discussion of each question.

**Conclusion:** This study identified several specific research questions generated by people living with chronic MSK pain, relatives, HCPs, and researchers. The stakeholders proposed prioritization of the healthcare system’s ability to support patients, focus on developing coherent pathways between sectors and education for both patients and HCP. These research questions can form the basis for future studies, funders, and be used to align research with end-users’ priorities

## Background

According to the World Health Organization (WHO), between 20-33% of the world population lives with a painful musculoskeletal (MSK) condition (1). As the costs due to MSK pain correspond to almost 2% of the gross domestic products of European countries, MSK pain poses a challenge for healthcare systems across the world (2,3). People with chronic MSK pain have a high use of healthcare services, reduced work ability, loss of productivity, and lower quality of life (4,5). In Denmark, it is estimated that 1.2 million Danish citizens (approximately 20% of the Danish population) experience chronic MSK pain some time during life (6). One in every four long-term sick leaves and 14% of all early retirements are caused by chronic MSK pain (6,7). Many people with chronic MSK pain have widespread complaints and deal with concurrent insomnia, anxiety, depression, and loneliness (8–11). This underlines that chronic MSK pain is a multifaceted problem and affects all aspects of life, limiting activities of daily living and work ability while increasing medical expenditure.

To identify the individually lived experiences with a chronic pain condition, there is a need to include their perspectives in the research agenda. The emerging concept of early and continuous involvement of people living with different conditions in research is internationally recognized as a priority (12,13). A recent example is the International Association for the Study of Pain (IASP) establishing the Global Alliance of Partners for Pain Advocacy (GAPPA). Previous research has shown that the traditional research does not always align with the needs and preferences of people living with a pain condition and healthcare professionals treating the condition (HCPs) (14).

To accommodate the lack of end-user involvement, the James Lind Alliance (JLA) was established to create a framework for Priority Setting Partnerships (PSPs) which provides a platform where end-users and researchers can work together on shaping an agenda for future research within a certain field. In the PSP framework, surveys and workshops are used to capture questions from end-users and to define research priorities that can address future research on replying unanswered so called uncertainties (17). Research priorities can vary across social groups and geographically between and within a country, highlighting the need for national initiatives to uncover uncertainties (18). Several studies have investigated research priorities within MSK pain either using other approaches (19–21) or in specific MSK conditions such as knee arthroplasty, spinal cord injuries, fibromyalgia (22–24), but no studies have investigated the research priorities in relation to a broader scope of chronic MSK pain within a national Danish context using the JLA-PSP approach. The objective of this study was to establish a list of top-research priorities for chronic MSK pain to formulate specific research questions using the JLA-PSP framework.

## Methods Study Design

This study is reported according to the Guidance for Reporting Involvement of Patients and the Public (GRIPP2) short form checklist (25). The study was conducted using a modified version of the JLA framework for conducting PSPs. It differed from the traditional approach by not using a specific JLA advisor as the aim was to develop specific research questions instead of producing a top-10 list. The methodology aimed at gathering inputs from several key stakeholders involved in the management of daily living with chronic MSK pain in Denmark, including people with lived experience, relatives, HCPs, and researchers. The study consisted of 4 phases, including: initial online survey, survey analysis, interim prioritisation exercise, and an online workshop. Additionally, the project group included three experts with previous experience conducting PSPs within chronic pain (KB, JS, and PP). All data were collected and managed through REDcap (Research Electronic Data Capture) hosted at Aalborg University (26,27). REDcap is a secure, web-based software platform designed to support data capture for research studies. The study was exempt from a full ethical approval by the North Denmark Ethical Committee due to the design of the study.

### Project Organisation

We established both a project group (PG) and a steering group (SG). The PG was in charge of developing the protocol, managing data, and publishing the results. The PG was restricted to researchers and research-HCPs. Additionally, we established both a SG restricted to people with lived experience and patient organisations and a SG restricted to HCPs, chaired by KDL. The SGs ensured equal input to all steps in the process both from people living with chronic MSK pain and HCPs. The SG included people with lived experience and representatives from three different patient organisations (the Association for Chronic Pain Patients, the Danish Fibromyalgia & Pain Association, and the Danish Rheumatism Association), researchers and HCPs working within the field of chronic MSK pain and at least one of the authors from the Danish national clinical guidelines relating to pain. The individually assigned tasks for the PG and SG can be seen in Table 1.

**INSERT TABLE 1 HERE – LEGEND: Definitions of Project and Steering Groups**

### Participants

For all steps of the PSP process, we included both people living with chronic MSK pain, relatives to people living with chronic MSK pain, HCPs, and researchers who were working with people living with chronic MSK pain. People living with chronic MSK pain were self-assessed based on the new definitions from the ICD-11 codes and the classification of chronic primary pain:

> *“Pain in one (or more anatomical regions) that persists or recurs for more than 3 months and is associated with significantly emotional distress or functional disability (interference with activities of daily life and participation in social roles) and that cannot be better accounted for by another chronic pain condition”* (28,29)

Relatives and people living with chronic MSK pain fulfilling the above were included. Health-care professionals were considered eligible for participation if they presently worked full-time with treating people living with chronic MSK pain. Also, only authorised HCPs including medical doctors, nurses, chiropractors, physiotherapists, occupational therapists, psychologists, or social workers were eligible to participate. Researchers were included if they had three years or more of experience within the field of chronic MSK pain and at least one first authored or senior authored paper within the field. People with chronic secondary pain (e.g., sequelae from chemotherapy, surgical procedures, or visceral pain) were not eligible for participation.

### Phase 1: National Online Survey including Multiple Stakeholders

To collect evidence of uncertainties in relation to chronic MSK pain in Denmark, we created a respondent-tailored online survey for people with lived experience, relatives, HCPs, and researchers. The survey was distributed using a multimodal recruitment strategy through personal networks, patient organisations, and social media, thereby covering all geographical areas of Denmark. The survey was developed within both SGs and in an iterative collaboration with people representing ethnical minorities in Denmark and linguists to ensure that the survey would be easily understandable, independent of reading capabilities. The survey included demographic questions, an option (i.e., “Which questions do you want future research to answer”) to capture the priorities from the respondents (a maximum of five questions were given) and finally, an invitation to participate in future steps of the process and/or in future projects in relation to the chronic MSK pain. None of the respondents were reimbursed for their participation.

### Phase 2: Analysis of Survey

The analysis of the survey data followed the step-by-step guidebook developed by JLA (17). Initially, data was imported into NVivo 12 for qualitative data analysis. Secondly, questions were sorted through naive reading and excluded if the questions were out-of-scope, unidentifiable, lacked information, or too individualised. Excluded questions were reviewed by at least two SG members, one person with lived experience and one representative from a patient organisation to ensure that the questions were truly out-of-scope (e.g., irrelevant to this area, not understandable). Thirdly, the remaining eligible questions were categorised in NVivo and sorted into initial themes, main themes, and underlying sub-themes by one author (KDL). Themes with < 5 individual uncertainties were excluded by the concept of saturation (30). Fourthly, indicative questions were formulated as true to the data as possible and rephrased into one theme when several questions fitted into one. The final list of indicative research questions was co-developed with both SGs to ensure relevance for all stakeholders.

### Phase 3: Interim Prioritisation

To reduce the final list of indicative research questions prior to the priority-setting workshop, we conducted an online interim prioritisation exercise. Participants from phase 1, who agreed to participate in further steps, were invited to take part in the interim prioritisation exercise. Participants were asked to rank the importance of each research question on a 5-point Likert scale from 1 to 5; with 1 = not at all important, 2 = low importance, 3 = Neutral, 4 = Important, to 5 = very important. The scores for all participants were merged and thus illustrated which research questions were deemed most important by the participants. These were included in the priority-setting workshop.

### Phase 4: Priority-Setting Workshop

A final list of the top research priorities was established by inviting people, relatives, and HCPs to participate in a 3-hour online workshop. Participants were recruited from the national survey or professional networks and were included on a first come, first served basis. The workshop used a nominal group technique approach with a combination of small and large group exercises, which were facilitated by two individuals from the project group (KDL and MSH) and an external facilitator, all with prior experience in conducting workshops. To establish the final list, we used a consensus approach and a final voting to choose the top priorities. All participants were split into three groups with representation of both persons with lived experience and HCP in every group. All questions were discussed and reviewed, and the least important research priorities determined by consensus were removed first. The following step focused on sorting and agreeing on the most important research priorities within the three groups before presenting for the entire group. Finally, all participants were asked to choose the two most important research priorities in their view which then constituted the final list of priorities.

## Results

### Phase 1: National Survey including Multiple Stakeholders

The survey was viewed by 1017 participants and completed by 1010 (89.5% people with lived experience, 8.8% HCPs, and 1.7% relatives) who suggested 3121 individual research questions. 64% of the HCPs worked in the primary sector, 35% worked in the secondary sector, and 37% worked in the tertiary sector. 675 respondents agreed to participate in later steps of the process (66%). Participants’ characteristics for phase 1, 3, and 4 can be seen in Table 2.

**INSERT TABLE 2 HERE – LEGEND: Participants’ Characteristics**

### Phase 2: Analysis of Survey

127 uncertainties (4.24%) were excluded for being out-of-scope, unidentifiable, or missing information. The remaining 2994 uncertainties were then transformed into 50 initial themes (See table 3) and then condensed into 19 main themes and 36 sub-themes which were formulated into research questions (See table 4).

**INSERT TABLE 3 HERE – LEGEND: Initial Themes**

**INSERT TABLE 4 HERE – LEGEND: Thematic Analysis**

### Phase 3: Interim Prioritisation

Participants from phase 1 and all members from both steering groups (n=19) were invited to participate in the interim prioritisation exercise and 97 people (45 people living with chronic MSK pain, 4 relatives, 34 HCPs, and 14 researchers) completed the exercise (24% completion rate). Based on the interim prioritisation exercise, the seven research questions with the lowest scores were excluded. This left us with 33 research questions, which were deemed an appropriate number of questions to handle at the priority-setting workshop (See Appendix 1).

### Phase 4: Priority-Setting Workshop

Twenty-three participants (including 11 people living with chronic MSK pain, 1 relative, 10 HCPs, and 1 researcher) participated in the online workshop to establish the most important research priorities in relation to chronic MSK pain. The final voting for the most important research questions can be seen in Table 5. After initial discussions in the group phase, all participants discussed and voted for the most important research questions related to chronic MSK pain. This prioritized list of research questions can be seen in table 5. Full list of research questions for the workshop can be found in Appendix 2.

**INSERT TABLE 5 HERE – LEGEND: Final End-user Generated Research Priorities for Chronic Musculoskeletal Pain**

## Discussion

Via an iterative process as a collaboration between people living with chronic MSK pain, relatives, HCPs, and researchers, we developed a set of research priorities for research areas of interest within the domain of chronic MSK pain in Denmark. Our process demonstrated that future research should prioritise investigation of how the healthcare system can offer support and better pathways between sectors, thereby leading to a coherent collaboration between patient, relatives, municipality, and various HCPs. Increased focus should also be given on education of patients, relatives, and HCPs to increase the general knowledge across all stakeholders involved in the management of chronic MSK pain. Overall, the PSP study has highlighted which research questions that the stakeholders within chronic MSK pain would like to see investigated.

### Comparisons with Previous Priority Setting Partnerships

The scope of our PSP was intentionally broad and included chronic MSK pain and not a specific diagnosis as e.g., hip or knee osteoarthritis. Our choice was based on current understanding that many people suffer from MSK pain in multiple sites where the pain and impact of pain are the most important aspects, and not the specific pathology. Patients with chronic MSK pain often present themselves with widespread symptoms, influenced by psychological and social factors (1,2). This highlights that chronic MSK pain is a multifactorial condition. Despite not being related to a specific condition, our PSP identified similar research priorities as several other PSPs which identified improvements of treatments for pain and function (e.g., knee arthroplasty) (22,31–35), improving health-care organisation (e.g., paediatric chronic pain) (22,33,34), improving the knowledge of living with a painful condition (e.g., people with fragile fractures in lower limb and pelvis) (31,33,34), improving the accuracy of diagnosis and referral (e.g., shoulder surgery (22,36,37) and improving quality of life and public awareness, and avoid stigmatisation as seen in other conditions such as depression, dementia, diabetes, and cancer (38–41). As a contrast to previous PSPs, our priorities included a strong focus on areas outside typical treatments focusing on improving pain and function. We identified priority areas that included an improved societal understanding and organisation of healthcare for people living with chronic MSK pain. The difference in priorities compared with previous PSPs may reflect a time-based change in the perception of chronic MSK pain. Previously considered a primarily biomedical condition, chronic MSK pain is now considered a condition developed from and maintained by multiple factors, which may co-exist with other conditions (3–5) (REFS). This highlights that to target these research priorities, it is essential to collaborate and form partnerships across institutions and sectors and involve researchers outside the healthcare profession.

### Participatory Research

Engaging people with lived experience has been endorsed by multiple organisations, including the WHO and IASP, and is seen as a crucial step in solving complex problems within society (42). Enabling the participation of relevant stakeholders in research provides a deeper understanding of real-life problems and facilitates better implementation of research findings (43,44). Additionally, co-creation of research helps mitigate research-waste while guiding decision-makers in changing policies within the health-care system (45). This study was an example of participatory research, which was co-created with relevant stakeholders to chronic MSK pain. While this approach has multiple advantages, there is still no consensus on the appropriate way to utilise and interpret findings from PSPs into real-life settings. Thus, our results should not be seen as a definitive list of research questions, but as a guide to research areas that reflect the problems faced by people living with chronic MSK pain as well as stakeholders.

### Limitations

To capture a broad large sample and limit regional differences, we used an online survey where all who fulfilled the criteria could respond to the survey. Ultimately, we ended up with a strong overrepresentation of female (93%), the majority were persons living with chronic MSK pain (90%), Danish (97%), and had been living with chronic MSK pain for more than 10 years (∼70% of people living with chronic MSK pain). It is unknown if this sample represents the broader population or if these individuals may have certain preferences and priorities. Despite attempts to engage participants with different ethnical backgrounds, we failed to recruit a substantial proportion of participants with other ethnicities than Danish. Minorities often are overlooked in policy making (46–49), and specific efforts are needed to engage these groups in future validation work of this PSP.

### Future Implications

The JLA-PSP approach recommends surveys for collecting research questions. This initial survey lays the foundation for the entire PSP. It might be argued that other approaches may be used to complement this approach as the lived experience of chronic MSK pain might be better understood using phenomenological approaches. Such an approach would provide unique insights into the perspectives of how people perceive their own life experiences with a certain condition, and how knowledge is gained from these experiences (50). Future studies could consider collecting research questions using qualitative methods such as interpretive phenomenological interviews or attempting to cross-validate the findings from this study using such methods.

Due to COVID-19, we were unable to host a physical workshop as recommended by the JLA (17). Therefore, we conducted an online workshop through Teams. It is unclear what effect such an approach may have. The evidence regarding the comparison between online and physical workshops is scarce and needs to be further investigated in future studies. Our experience with online workshops were predominately good, and it allowed people to participate without challenges with transportation, needing to go outside of your home, and limiting time requirements for busy participants. Furthermore, this medium made it easier and safer for more vulnerable participants to feel comfortable. Future research is needed to understand if these online workshops may be a new tool to engage groups which have earlier been underrepresented in PSPs.

## Conclusion

While our knowledge on the complexity of the lived experience with chronic MSK pain has increased during the past decades, there are still several unanswered questions. Findings from this PSP can guide researchers and funders to prioritise specific research areas within chronic MSK pain, thereby potentially leading to important improvements within the management of chronic MSK pain. Using the JLA-PSP method and engaging stakeholders in all aspects of the study, we were able to retrieve a list of research priorities from people living with MSK pain, relatives, and healthcare practitioners working with people with chronic MSK pain. To be able to target these research priorities, cross-sectoral collaboration, partnership between stakeholders, and willingness to engage in establishing coherent pathways are required. Our research group has initiated activities to uncover some of these research questions, and we hope that other institutions will also engage in stakeholder driven research activities in future.

## Data Availability

All data produced in the present work are contained in the manuscript.

