## Supplementary material for "Participatory Research: A Priority Setting Partnership for Chronic Musculoskeletal Pain in Denmark": Tables, Figures and Appendix 1-2

**Tables and Figures (Research Priorities)**

***Table 1. Definitions of project and steering groups***

| **What** | **Who** | **Role and Responsibilities** |
| --- | --- | --- |
| Project group | Researchers and research-clinicians | Protocol preparation, study management, data management and analysis, publication, knowledge translation. |
| Steering Groups | People with CMP, their relatives, relevant patient-organizations, HCPs, and researchers. | Providing inputs to protocol, survey, categorization, interim ranking, workshop, final formulation of research questions and knowledge mobilisation |

Note: HCP = Health-care practitioners; CMP = Chronic musculoskeletal pain

***Table 1. Definitions of project and steering groups***

|  | **No. (%) of participants** | | |
| --- | --- | --- | --- |
| **Table 2. Participant Characteristics** | Phase 1: Survey *n* = 1010 (%) | Phase 2: Interim prioritisation *n* = 97 (%) | Phase 3: Workshop *n* = 23 (%) |
| **Participants** |  |  |  |
| Person living with chronic MSK pain | 904 (89.50) | 45 (46.39) | 11 (47.82) |
| Relative to a person living with chronic MSK pain | 16 (1.58) | 4 (4.12) | 1 (4.34) |
| Medical Doctor | 15 (1.48) | 13 (13.40) | 2 (8.69) |
| Physiotherapist | 22 (2.21) | 15 (15.46) | 6 (26.08) |
| Other HCPs (Psychologist, Nurse, Chiropractor, Social worker, researcher) | 53 (5.22) | 20 (20.61) | 6 (26.04) |
| **Sex** |  |  |  |
| Female | 939 (92.97) | 75 (77.3) | 21 (91.3) |
| Male | 71 (7.03) | 22 (22.7) | 2 (8.7) |
| **Age** |  |  |  |
| 18-30 | 52 (5.1) | 3 (3.0) | 1 (4.3) |
| 31-40 | 106 (10.4) | 44 (47.4) | 3 (13.2) |
| 41-50 | 283 (27.9) | 12 (12.4) | 12 (52.2) |
| 51-60 | 366 (36.0) | 25 (25.7) | 6 (26) |
| 61-70 | 168 (16.5) | 12 (12.4) | 1 (4.3) |
| 71-80 | 39 (3.8) | 0 | 0 |
| 80+ | 2 (0.2) | 0 | 0 |
| **Region** |  |  |  |
| Region of Northern Jutland | 130 (12.8) | 27 (27.8) | 6 (26) |
| Region Central Denmark | 227 (22.3) | 25 (25.7) | 4 (17.3) |
| Region Southern Denmark | 189 (18.6) | 18 (18.5) | 3 (13.2) |
| Region Zealand | 206 (20.3) | 12 (12.4) | 6 (26) |
| Region Capital | 264 (26) | 15 (15.4) | 4 (17.3) |
| **Ethnicity** |  |  |  |
| Danish | 979 (96.4) | 94 (97) | 22 (95.7) |
| Immigrant, non-western | 7 (0.7) | 0 | 0 |
| Immigrant, western | 15 (1.5) | 1 (1) | 1 (4.3) |
| Descendant, non-western | 2 (0.2) | 1 (1) | 0 |
| Descendant, western | 1 (0.1) | 1 (1) | 0 |
| Do not want to state | 12 (1.2) | 0 | 0 |

***Table 2. Participant Characteristics***

**Table 3:**

**Initial Themes**

Biomedical approach, Biopsychosocial approach, Care-pathway, Characteristics, Classification, Cognition, Coping, Cross-sectional Management, Diagnosis, Education (Clinicians), Education (People living with chronic MSK pain), Environment, Equal treatment options, Ethnicity, Fatigue, Financial support, Gender differences, Genetics, Implementation, Insurance, Job centre, Measurement, Mechanism, Municipal management, Needs and preferences, Nudging, Nursing, Nutrition, Pacing, Pain Fluctuations, Pain Spreading, Physical limitations, Prevention, Prognosis, Psychological, Public Awareness, QoL, Relatives, Self-management, Sequalae, Sex, Shared decision-making, Sleep, Stigma, Stress, Therapeutic alliance, Treatment, Understanding of pain, Vitamins, Work.

***Table 3. Initial Themes***

***Table 4. Thematic Analysis of Survey***

| **Main theme** | **Sub-theme** | **Indicative question [uncertainty]** | **Example of original uncertainty submitted** | **No. uncertainties** | **Summary** |
| --- | --- | --- | --- | --- | --- |
| **Care-pathway** | Retention | How can the healthcare system avoid ‘losing’ people living with chronic MSK pain dealing with CMP, and keep them in the ‘loop’ independent of income and geography? | *Hvordan undgår sundhedssystemet at tabe en borger med komplekse universelle slidproblematikker i alle led på gulvet/imellem stole?* | **41 People living with chronic MSK pain** | This question mainly focuses on how people living with chronic MSK pain that deal with severe disability, who often experience that they have been forgotten by the healthcare system, and therefore needs research on how the system can better retain people living with chronic MSK pain who needs to be retained. |
| **Care-pathway** | Referral | How can we ensure that people living with chronic MSK pain receive the appropriate referrals, quicker? | *Hvorfor er det så svært og få henvisninger til speciallæger når man har mange kroniske smerter?* | **66 People living with chronic MSK pain** | N/A |
| **Care-pathway** | Flow | How can current care-pathways be improved to ensure a better more fluid organisation and thereby create a better flow in care-pathway with shorter waiting-times? | *Hvordan kan man organisere sundhedsvæsenet så patienten ikke bliver en kastebold, og således kan udredes / blive diagnosticeret og evt. smertebehandlet på kortere tid?* | **92**  **All groups** | This question derives from uncertainties relating to care-pathways that is often considered as inefficient and therefore associated with long periods of uncertainty. |
| **Care-pathway** | Cross-professional | How can specialised pain clinics support people living with chronic MSK pain living with CMP and can it be a better individualised alternative to usual care-pathways? | *Hvordan kan vi gøre udredning af sammensatte sygdomme hurtigere og med input fra flere specialer? Hvordan kan vi se "elefanten" i sin helhed og ikke blot et ben ad gangen?*  *_________*  *Hvilke behandlingsstrategier resulterer i de bedste patientoplevelser i forhold til smertehåndtering.? Eksempelvis enkeltekspert-behandling vs smerteteams (med læge, psykolog, fysioterapeut, socialrådgiver, ergoterapeut)* | **108 All groups** | This question evolves around the need for improved care-pathways and as an alternative to usual care, all groups want to know how cross-professional management, for example using specialized pain clinics, can be used as potential ‘better’ care pathway for people living with chronic MSK pain with CMP |
| **Care-pathway** | Coordinator | What would be the effect of having support-staff to coordinate rehabilitation of people living with chronic MSK pain living with CMP? | *Kunne der være en fordel i at have en koordinator tilknyttet så patienten ikke er tovholder i eget smerteforløb?* | **29**  **People living with chronic MSK pain** | People living with chronic MSK pain with CMP who lacks the greater overview over the different offers that exist within the healthcare system, wants to know how personalised help from a coordinator (i.e. staff) can improve their experience with their care-pathway. |
| **Complications** | Cognition | How does CMP affect the patient in pain and how can we decrease the impact that pain has on cognition, concentration and memory? | *Kroniske sygdomme påvirker ikke kun krop, men også hjernen med kognitive dysfunktioner, hvordan får man behandling heraf integreret - fx. træning af hukommelse?* | **28 People living with chronic MSK pain** | N/A |
| **Complications** | Symptoms | How can we explain the symptoms that people living with chronic MSK pain with CMP experience and how can we explain the changes (e.g. fluctuations, spreading) that occur in people living with chronic MSK pain with CMP? | *Hvorfor er smerten varierende i styrke og springer rundt i kroppen* | **31 People living with chronic MSK pain** | N/A |
| **Complications** | Fatigue | How common is fatigue in people living with chronic MSK pain with CMP, why do people living with chronic MSK pain feel fatigued (mechanism) and how do we decrease the levels of fatigue in people living with chronic MSK pain with CMP (treatment)? | *Hvordan man kan modvirke udmattelsen der ofte medfølger smerte sygdomme, både i hovedet i form af hjernetåge og i hele kroppen i form af muskeludmattelse.* | **45 People living with chronic MSK pain** | N/A |
| **Cross-sectional Management** | Collaboration | Which effect does cross-sectional management of people living with chronic MSK pain with CMP have compared to usual care and if more effective, how do we improve current cross-sectional management? | *Hvordan sikres en sammenhængende tværfaglig behandling samt samarbejde med de kommunale instanser bedst?* | **77**  **All groups** | Based on the uncertainties from the survey all groups question the inefficient or in some cases absent cross-sectional management and therefore, asks how cross-sectional communication and general management can be improved to further improve the life of people living with chronic MSK pain with CMP. |
| **Diagnosis** | Diagnostic uncertainty | How does diagnostic uncertainty impact people living with chronic MSK pain with CMP and how can (earlier) diagnostic clarity improve acceptance of life circumstances? | *Mange patienter går rundt uden diagnosticering. Hjælper diagnosticering patienter med accept og afklaring af ændrede livsvilkår?* | **55 People living with chronic MSK pain** | This theme revolves around how we can improve the way clinicians diagnose CMP and how this can be performed faster, more accurate and how we can identify those people living with chronic MSK pain predisposed for CMP.  Lastly, people living with chronic MSK pain want research that uncover the impact of diagnostic uncertainty and how this uncertainty can be dealt with the help of the healthcare system. |
| **Diagnosis** | Fast-track | How can we reduce the time to diagnosis and minimise errors in diagnosing people living with chronic MSK pain with CMP? | *Hvordan får man stillet ind hurtigere diagnose uden at man skal være kastebold ved flere læger? Kan man ikke få en blodprøve eller lignende, så man undgår de fejl, som gør at man ikke får en diagnose og bliver set skævt på?* | **27 All groups** |  |
| **Diagnosis** | Test | Why is it so difficult to diagnose CMP and how can we improve test to diagnose CMP more efficiently? | *Hvorfor er det så svært at få en konkret diagnose? – jeg har været ved utallige behandlere og det har taget mig over 10 år at få en endelig diagnose*  *_____ Kan man påvise fibromyalgi på en anden måde end kun tenderpoint test og sygdomsforløb? F.eks. vha. en blodprøve så diagnosen bliver fuldstændig sikker. Og kan man måle det præcise smerteniveau helt objektivt, så det ikke blot bliver en personlig karakter fra 1-10?* | **40 People living with chronic MSK pain** |  |
| **Diagnosis** | Early identification | How can we identify/trace those people living with chronic MSK pain predisposed for CMP or diagnose people living with chronic MSK pain earlier (i.e. teenage years)? | *Hvordan havde mit smerteforløb set ud hvis det var blevet opdaget tidligere? Kan det forhindre mine børn i at få det?* | **31 People living with chronic MSK pain** |  |
| **Education (People living with chronic MSK pain)** | Empowerment | How can patient education be improved in order to make patient more knowledgeable in their own condition and thereby take more ownership and self-manage? | *Hvordan får man informeret og empowered patienterne om deres videre forløb i livet så de selv kan gøre noget ved smerterne?* | **45**  **All groups** | All groups gave survey responses which led to the questions, 1) what do patient educational packages need to entail and how should they be delivered and 2) how can we improve the education the people living with chronic MSK pain receive and how can it empower the people living with chronic MSK pain’ self-management. |
| **Education (People living with chronic MSK pain)** | Delivery | What do people living with chronic MSK pain living with CMP need to know and how should patient education be delivered? | *Hvad er patient uddannelse og hvilke emner i patientundervisning har størst effekt på patientens oplevelse af smertestabilisering/smertenedgang?* | **21**  **All groups** |  |
| **Education (HCP)** | Knowledge | What do clinicians need to know and how can we improve the general level of knowledge of CMP ensure better management of these people living with chronic MSK pain? | *Hvad er viden og uddannelsesniveauet indenfor forskellige faggrupper og hvor stor er denne forskel på tværs af institutioner i landet?* | **180**  **All groups** | This theme asks the questions 1) what do clinicians need to know about CMP and 2) how can the knowledge of HCPs be increase in order to improve general care of people living with chronic MSK pain with CMP. |
| **Mechanism** | Mechanism and  risk factors | What is the mechanism and which risk factors (e.g. other illness) are associated with developing CMP? | *Hvilke fysiologiske faktorer, fx relevant absolut styrke, lednær-muskelmasse, BMI, balance (neuromuskulær kontrol) eller billedediagnostik (graden af artrose) osv. er stærkest associeret med smerter og livskvalitet?*  *_____*  *Hvorfor får man kroniske smerter? Hvad er årsagen til mine smerter?* | **560**  **All groups** | This theme asks how 1) CMP develops in persons, 2) who’s at risk for developing it and which risk factors and 3) how does common treatment options work. |
| **Mechanism** | Gender differences | Which role does gender plays in developing CMP and how can this knowledge guide management? | *Hvorfor er der forskel på kvinder og mænd der udvikler kroniske smerter?* | **12  People living with chronic MSK pain** |  |
| **Mechanism** | Treatment | What is the mechanism of action of the most common treatments and how can this knowledge improve the management of CMP? | *Hvad er den egentlige virkningsmekanisme for træning til kroniske smerte patienter? Hvorfor skal vi ikke bare give dem en pille mod smerterne?* | **315 All groups** |  |
| **Mechanism** | Genetics | What is the role of genetics in both developing pain and inherit pain? | *Er der forskelle i smertebehandlings-resultater mellem smerte-patienter, der har fået konstateret tilstanden, som den "første i slægten" og dem, som har "ned-arvet" den gennem flere generationer?* | **45 All groups** | This question revolves around people living with chronic MSK pain need to understand the role of genetics (i.e. epigenetics) and how genetics can be used to treat and understand pain, but also if CMP can be inherited to the next generation. |
| **Municipal Management** | N/A | How can we ensure that patient living with CMP receives the same support in the municipalities to avoid stigmatisation and unsatisfaction? | *Hvordan kan vi ensrette behandlingen af patienter med kroniske smerter og hvordan får man dem til at acceptere og anerkende vores smerter?* | **51**  **People living with chronic MSK pain** | This question revolves around how some people living with chronic MSK pain experience poor management when entering the municipalities and therefore, wants future research to focus on how to improve the management of people living with chronic MSK pain with CMP in the municipalities. |
| **Municipal Management** | Responsibility | What is the role and responsibilities of the municipalities in managing people living with chronic MSK pain with CMP and how can we improve the management that people living with chronic MSK pain receive in the municipalities? | *Hvordan indvirker kommunernes ansvarsområde på, hvordan afklaringsprocessen for en smerteramt bliver?* | **21**  **All groups** | N/A |
| **Nutrition** | Role | Which role does nutrition (including vitamins) have in the development and persistence of CMP? | *Er der sammenhæng mellem kostindtagelse og smerter i fibromyalgi. Er der fødevarer, der påviseligt direkte forværrer smerterne?* | **7**  **People living with chronic MSK pain** | N/A |
| **Nutrition** | Effect | What is the effect of various diets, foods and supplements on CMP? | *Hvor stor en rolle spiller kost og kosttilskud ift. behandling af fibromyalgi og forbedring af smertetilstanden?* | **38 All groups** | N/A |
| **Prognosis** | N/A | Which role does increases in pain play in the prognosis og pain, and how can we collect and disseminate knowledge on the prognosis to people living with chronic MSK pain living with CMP and what can people living with chronic MSK pain expect their lives to look like? | *Kan man risikere at forværre tilstanden på den lange bane, hvis man insisterer på at fastholde så normal en hverdag som muligt med arbejde, motion etc.?* | **78 People living with chronic MSK pain** | People living with chronic MSK pain want future research to focus on the knowledge of prognosis. More specific they need research to focus on how the action of todays impact their future daily life and if their pain condition can be improved or even completely resolve. |
| **Psycho-social** | Influence | What is the psycho-social consequences of dealing with CMP and what is the association between psycho-social factors and developing CMP? | *Hvad er sammenhængen mellem fx depression og da jeg udviklede kroniske smerter?* | **18**  **People living with chronic MSK pain** | This theme covers the need for future research to focus on the psycho-social impact on the life of people living with chronic MSK pain living with CMP. Derived from the uncertainties the questions that needs answered is 1) how psychological conditions influence CMP, 2) what impact does CMP have on both your psychological and social state and 3) Can interventions that specifically targets psychological and social aspects improve the lives of people living with chronic MSK pain with CMP. |
| **Psycho-social** | Management | What is to role of early psycho-social intervention in reducing pain and the impact of CMP on daily living? | *Hvordan organiseres behandling af sårbare patienter, der udover langvarige smerter også har psykiatriske diagnoser og/eller iatrogent misbrug. Denne gruppe af patienter falder ofte mellem flere stole, fordi hverken smertecentre eller psykiatri er klædt på til at varetage patienternes udfordringer, og der er ikke tradition for et samarbejde.* | **52**  **All groups** |  |
| **Quality of Life** | N/A | How does CMP impact QoL and how can different management strategies improve daily living? | *Jeg ved at jeg skal leve med mine smerter resten af mit liv, men hvordan kommer jeg til at få det lettere i hverdagen, hvordan kan systemet hjælpe mig at med kunne gøre mere i min hverdag?* | **38**  **All groups** | N/A |
| **Relatives** | Inclusion | How can we include relatives more into the situation of living with pain – how can we educate them and make them comfortable with their role living with a person with CMP? | *Fokus på håndtering af pårørende og nye bekendtskaber. Giv os hjælp og vejledning til at introducere andre til kroniske smerter. Til at få vores pårørende til at forstå hvor invaliderende sygdommen er og hvorfor vi ikke "bare kan gøre det".* | **10 People living with chronic MSK pain and relatives** | N/A |
| **Sex** | Dysfunction | How can the sexual life of people living with chronic MSK pain living with CMP be improved? | *Hvordan kan et intimt liv blive bedre for borgere med kroniske smerter i muskler og led, området er stærkt nedprioriteret?* | **8**  **People living with chronic MSK pain** | N/A |
| **Sleep** | Impact | Which consequences do limited sleep have on people living with chronic MSK pain living with CMP? | *Hvad er langtidsvirkningen af dårlig søvn – hvordan påvirker det mine smerter og overskud i løbet af dagen?* | **16**  **People living with chronic MSK pain** | With the help of people living with chronic MSK pain this theme asks, 1) how does limited sleep and/or poor sleep quality impact people living with chronic MSK pain living the CMP and 2) how can limited sleep and/or poor sleep quality be improved with intervention? |
| **Sleep** | Improving | What can we do to improve sleep quality in people living with chronic MSK pain living with CMP both using medical treatments and using natural treatments? | *Hvordan søvnproblemer bedre kan løses så hverdagen for patienten kan blive mere til at holde ud og der ikke er så store svinger i hverdagen eller i timerne på dagen* | **68 People living with chronic MSK pain** |  |
| **Stigmatisation** | Management | How severe is stigmatisation in current care and how does it influence the management of people living with chronic MSK pain with CMP? | *Findes der fordomme hos sundhedsprofessionelle om kroniske smerter? Kan det give dårligere eller fejlagtig behandling?* | **12**  **People living with chronic MSK pain** | People living with chronic MSK pain want to uncover the stigmatisation in clinical practice and how this influence decisions, treatment and the general management of clinicians throughout the care pathway in regard to people living with chronic MSK pain with CMP. |
| **Stigmatisation** | Diagnosis | How can increase awareness of the diagnosis as an equal diagnosis to other more ‘accepted’ conditions in order to reduce the misbelief that people living with chronic MSK pain with CMP often experience from the healthcare system? | *En afklaring af hvordan kroniske smerter skal rubriceres i sygdomskategori og inden for behandlingssystemet. Nogle regner det fortsat fejlagtigt som en form for psykisk sygdom, så man bliver tit afvist hos praktiserende læger, der ikke anerkender sygdommen som fysisk.* | **78**  **People living with chronic MSK pain** | Patient want future research to focus on the need for greater levels of awareness and acceptance of CMP within the healthcare system, so that the people living with chronic MSK pain don’t feel stigmatised when dealing with the healthcare system. |
| **Stigmatisation** | Public Awareness | How can we improve the knowledge of CMP in the public and how can increases public knowledge help people living with chronic MSK pain live a better life with CMP? | *Hvordan man bedre kan vise pårørende, omverden, arbejdspladsen osv. at fibromyalgi er en virkelig sygdom og ikke bare noget folk bilder sig og lader sig om?* | **34**  **People living with chronic MSK pain** | From the questions given be people living with chronic MSK pain this question revolves around the need to further increase the public awareness and knowledge in regards of CMP with the aim of reducing stigmatisation. |
| **Treatment** | Effectiveness | What is the most effective treatment option for people living with chronic MSK pain with CMP and how can the best treatment option be identified, individualised and delivered? | *Hvordan kan vi komme væk fra at behandle udelukkende på symptomer? Jeg vil gerne vide hvilken behandling der er bedst for mig og min sygdom?* | **903**  **All groups** | This question builds on the majority of responses from all groups and focusses on the need for developing better treatments or determining the most effective treatment for treating people living with chronic MSK pain with CMP. Furthermore, it revolves around how we can improve how we use existing treatments. |
| **Treatment** | Cost | What is the most cost-effective treatment to people living with chronic MSK pain with CMP and how can this knowledge be implemented into clinical practice? | *Hvilken behandling er mest effektiv både på smerter og økonomisk?*  *______ Forskningen bør identificere hvor der eksisterer forbedringspotentiale for "mere sundhed for pengene". Dette handler om a) at identificere uopfyldte behov og ønsker hos patienter og b) identificere de bedste indsatser* | **37**  **HCPs** |  |
| **Treatment** | Equality | How can we ensure that all people living with chronic MSK pain across age, population, ethnicity, health literacy and more will be offered the same treatment options? | *Forskningen bør identificere forbedringspotentiale for at reducere ulighed i sundhed. Dette handler om a) at identificere ulighed målt på forskellige parametre og b) identificere de mest effektive indsatser til at reducere ulighed* | **45**  **All groups** | Based on responses from both groups, this theme focusses on how we can ensure that treatments will be delivered to all people living with chronic MSK pain equally despite differences in demographics and more. |
| **Work** | Retention | How do you keep people living with chronic MSK pain living with CMP to continue working full time or part time and how can we make it easier to deal with work while living with pain? | *Hvordan vil man sikre hurtigere og bedre fastholdelse af smertepatienter arbejdsmarkedet?* | **38**  **People living with chronic MSK pain** | N/A |

**Table 5: Final end-user generated research priorities for chronic musculoskeletal pain**

| **Final prioritisation of research questions** | **Votes (n)** |
| --- | --- |
| How can specialised pain clinics support people living with chronic MSK paincompared to usual care-pathways? | 8 |
| How can we minimise errors in diagnosing people living with chronic MSK pain? | 7 |
| How can patient education be improved in order to make patient more knowledgeable in their own condition? | 6 |
| What is the most effective treatment option(s) for people living with chronic MSK pain? | 4 |
| How can we improve the general level of knowledge chronic MSK pain for clinicians to ensure better management of these people living with chronic MSK pain? | 3 |
| How can current care-pathways be improved to ensure a more coherent organisation? | 3 |
| How can we ensure that people living with chronic MSK pain receives the same support in the municipalities to avoid stigmatisation and dissatisfaction? | 1 |
| What is the mechanism and which risk factors (e.g. other illness) are associated with developing chronic MSK pain? | 1 |
| How can we improve the management that people living with chronic MSK pain receive in the municipalities? | 1 |
| What is the most cost-effective treatment to people living with chronic MSK pain? | 1 |

***Table 5. Final end-user generated research priorities for chronic musculoskeletal pain***

**Appendix 1**

**Appendix 1: Least important research questions**

**Excluded research questions based on interim prioritisation**

- What role does gender plays in developing pain and how can this knowledge guide management?
- What would be the effect of having support-staff to coordinate rehabilitation of people living with chronic MSK pain?
- What is the role of genetics in both developing pain and inheriting pain?
- How can the sexual life of people living with chronic MSK pain be improved?
- What role does nutrition (including vitamins) have in the development and persistence of chronic MSK pain?
- What is the effect of various diets, foods, and supplements on chronic MSK pain?
- What role does increases in pain play in the prognosis of pain, and how can we collect and disseminate knowledge on the prognosis to people living with chronic MSK pain and what can they expect their lives to look like?

**Appendix 2: Full list of research questions for the workshop**

**Research Questions**

What effect does cross-sectoral handling of people living with chronic MSK pain have, is it more efficient than ordinary handling and can we then optimize the cross-sectoral handling?

How can we reduce the time before getting a diagnosis while minimizing diagnostic errors?

How can patient education be improved so that people living with chronic MSK pain become better at dealing with their own pain and gaining more knowledge about their own condition?

What is the most effective treatment for people living with chronic MSK pain and how can this treatment be identified, individualized, and delivered in the best possible way?

What do clinicians need to know about chronic MSK pain and how can we improve the overall level of knowledge?

How can current patient processes be improved so that we create a more fluid process with shorter waiting times?

How can we ensure that people living with chronic MSK pain for a uniform treatment (municipal) and avoid stigma?

What is the reason for developing chronic MSK pain and what risk factors exist for the development of chronic MSK pain?

What role does your municipal management play in the treatment of people living with chronic MSK pain and how can we improve this management?

What is the most cost-effective treatment for people living with chronic MSK pain and how can this knowledge be implemented in practice?

How can we more quickly identify the people who are predisposed to chronic MSK pain?

How to keep people living with chronic MSK pain at work and how can we make it easier for people living with chronic MSK pain to work?

How can we reduce or prevent fatigue in people living with chronic MSK pain?

How can specialized pain clinics help people living with chronic MSK pain and is it a better alternative to regular management?

How can the healthcare system avoid losing people living with chronic MSK pain living and instead retain those who need it, regardless of income and geography?

How do we ensure that people living with chronic MSK pain receive the right referral, faster?

How can current care pathways be improved so that we create a more fluid process with shorter waiting times?

How can specialized pain clinics help people living with chronic MSK pain and is it a better alternative to regular management?

How can we improve diagnostic methods to provide a diagnosis more quickly and accurately?

What do people living with chronic MSK pain need to know about their condition and how should this knowledge be delivered?

What is the mechanism for developing chronic MSK pain and what risk factors exist for the development of chronic MSK pain?

How do respective treatments work and how can we use this knowledge to improve the treatment of chronic MSK pain?

What role does your municipal management play in the treatment of people living with chronic MSK pain and how can we improve this management?

What are the psychosocial consequences of living with chronic MSK pain and what role do psychosocial factors (eg anxiety, depression) play in developing chronic MSK pain?

How can we include relatives more and how can we educate them for more security?

What are the consequences of restricting sleep in people living with chronic MSK pain?

How can we improve sleep quality in people living with chronic MSK pain?

How can we improve knowledge about chronic MSK pain in the public and how can this knowledge affect the lives of people living with chronic MSK pain?

How can we ensure that all people living with chronic MSK pain, regardless of background, are offered the same treatment and thereby create a more equal healthcare system?
